# COVID-19 trend in Bangladesh: deviation from epidemiological model and critical analysis of the possible factors

**DOI:** 10.1101/2020.05.31.20118745

**Authors:** Asif Ahmed, Mohammad Mahmudur Rahman

**Author notes:** **Correspondence: Asif AHMED**. **Ethics approval and consent to participate:** not applicable. **Availability of data and material:** Secondary data used and cited in this study are publicly available. **Competing interests**: Authors declare no competing interests. **Authors’ contributions**: AA conceived the idea; AA and MMR equally contributed to the manuscript and agreed to submit in its current form after necessary corrections.

## Abstract

**Background:** Since its first report on March 08, COVID-19 positive cases and number of deaths are increasing in Bangladesh. In the first month of COVID-19 infection, incidence of daily positive cases did follow the susceptible, infected and recovered (SIR) based predictions we reported in April, but started to deviate in the following months. COVID-19 transmission and disease progression depends on multifaceted determinants e.g. viral genetics, host immunity, social distancing, co-morbidity, socio-demographic and environmental parameters. Therefore deviation in confirmed cases from predicted model may appear and warrant thorough investigation.

**Methods:** In this short report, we compared real data with SIR model and analyzed the possible factors associated with the deviation which included preventive intervention strategies, socioeconomic capabilities, climatic and meteorological indexes, acquired immunity of Bangladeshi population, demographic characteristics, health indicators and food habits.

**Results:** The key factor responsible for the observed deviation was found to be the number of tests performed. Having population with low median age, young age groups are being mostly infected. Low prevalence of non-communicable diseases among them and strong immunity compared to the elderly might have kept most of them asymptomatic with silent recovery. Warm temperature, humidity and UV index of Bangladesh during this summer period might have contributed to the slow progression of infection. Longer daylight mediated immunity, fresh air circulations and ventilation, less population density in rural areas and certain food habits perhaps helped the large number of populations to restrict the infection up to a level.

**Conclusion:** Despite all these helpful determinants in Bangladesh, person to person contact is still the leading risk factor for COVID-19 transmission. Infection may increase rapidly if safe distance and preventive measures are not strictly followed while resuming the normal social and work life. Expanding test capacity, strong collaborative action plans, strategies and implementation are needed immediately to prevent catastrophe.

**Highlights:**

1. Limited number of tests compared to large population was the key reason for possible low daily positive cases reported in Bangladesh.
2. Controlled interventions viz. official leave; transport ban and social distancing had helped initially to slow down the transmission.
3. Warm weather, high humidity and UV index, sunlight mediated immunity, fresh air circulations, low pollutions, food habit and heterologous immunity might have reduced the transmission capabilities of SARS-CoV-2.
4. Having large number of young people with strong immunity might have kept most of the infected asymptomatic who recovered silently.
5. Person to person contact still remain as key risk factor in COVID-19 transmission, so strict health measures should be in place even after reopening social activities to contain further transmission.

## 1 Introduction

Bangladesh is still experiencing daily rise of COVID-19 cases and deaths since its first report on 08 March, 2020. As of 22 June 2020, a total of 1,15,786 confirmed cases, 46,755 recovery and 1,545 deaths have been recorded [1]. After its first incidence in Wuhan, China in December 2019, COVID-19 spread to most of countries mostly via international travelers. Bangladesh government initially started with 10 day travel ban across the country from 26 March along with office and educational institutional shut down which was further extended [2]. People were advised to stay home and to maintain social distance, however it was difficult for daily wage earners and for the people staying in very dense premises [3]. During official leave and transport ban some unexpected mass gathering took place including back and forth movement of garments workers to Dhaka, large funeral prayer and crowded journey toward rural areas [1]. But overall the progression of COVID-19 in Bangladesh seemed to be relatively slow.

Susceptible, infected and recovered (SIR) is a classic closed compartment epidemiological model to predict disease trajectory [4]. After the rapid spread of COVID-19 many SIR and modified SIR based prediction have been reported [5-8] and classic SIR model is claimed to show less complexity and better prediction compared to modified models [9]. These predictions are not always perfectly correct as disease progression is determined by many other factors, however, such predictions help to visualize possible intervention mediated outcomes and action planning can be directed accordingly. In our previous report, we used SIR based prediction of COVID-19 in Bangladesh with different percentage of possible social distancing intervention [5]. Based on another survey and practical scenarios we hypothesized around 60% social distancing could be possible in Bangladesh [10]. In accordance, we showed COVID-19 in Bangladesh might reach its peak in early June and would be slowed down at the end of August. The daily cumulative case number was following our prediction graph very closely till late April. However, after that point confirmed cases did not follow the prediction trend and fell way behind it. As COVID-19 incidence is being controlled by multifaceted parameters, we tried to connect the possible reasons to the ongoing trend of Bangladesh. We considered demographic and climatic parameters, logistic and intervention strategies taken by Bangladesh in coordination with published literature to support the possible reasons.

In this short report, we investigated the deviation pattern of COVID-19 cases from our previously reported SIR prediction model of Bangladesh. We also tried to explain possible reasons behind this deviation and limiting factors in the light of literature. We followed some key events in Bangladesh during this period as well as environmental and behavioral pattern that might have influenced the outcome. We also tried to predict probable incidence number if increased number of tests could be performed. In harmony with global scientific data and suggestions the present and future risk factors in Bangladesh were highlighted which must be addressed to limit rapid progression of COVID-19.

## 2 Methods

COVID-19 related data of Bangladesh viz. total test numbers, total positive cases, total deaths and total recovery were retrieved on June 22, 2020 [1]. Test positive rate (%), daily percent change in test numbers and daily percent change in positive cases were calculated and plotted. Daily total positive case numbers were further adjusted subtracting deaths and deaths plus recovery. Findings were compared with our previously reported SIR model and trend of related parameters were analyzed. Global COVID-19 data were retrieved from Worldometers on 22 June, 2020 [11]. GraphPad Prism v. 6.0 was used for plotting and statistical analysis was done with linear regression.

## 3 Results

### 3.1 Comparison of reported data and SIR model

We compared previously reported SIR prediction data with real COVID-19 positive data of Bangladesh from the last three months. It was observed that real cumulative cases closely followed the prediction model in the initial month but later deviated from the predicted numbers (Figure 1A and 1B). The predicted model with consideration of about 60% population under lockdown had its peak on June 06; however the real case numbers are way below the prediction and still showing the rising trend shown in inset (Figure 1B).

**Figure 1:**
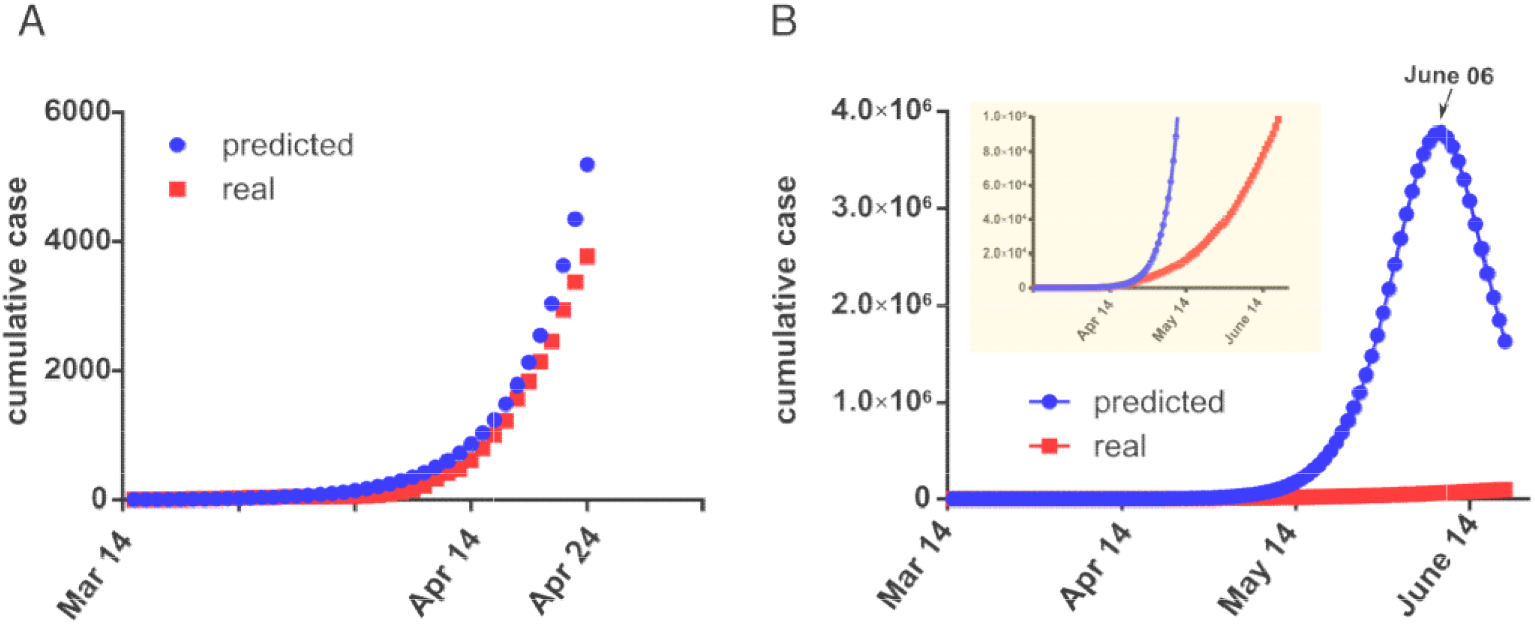
Graphical representation of COVID-19 SIR model and deviation in real data in Bangladesh. (A), previously reported SIR model with predicted and real cumulative cases from March 14 to April 24, 2020 (adapted from [5]); (B), same model compared extended up to June 21, 2020; the inset shows trend of real cases with zoomed y axis data.

We first looked into the relationship between daily test number and positive cases. The daily test number and positive cases showed linear correlation (R^2^=0.9601) over the entire period (Figure 2A). Until midway positive cases were below the straight line, however with increasing positive cases data tend to shift from the straight line as time progressed. We also observed that in the first month although data jumped around a bit, daily percent increment in test number and positive cases went hand in hand afterwards (red and blue line) (Figure 2B). From the beginning of May, as new number of tests got limited, slow decline of these two values were observed when test positive (%) were found to be increasing (Figure 2B). During this period the test positive (%) increased from around 10% to 20% and remained almost stable in final two weeks.

**Figure 2:**
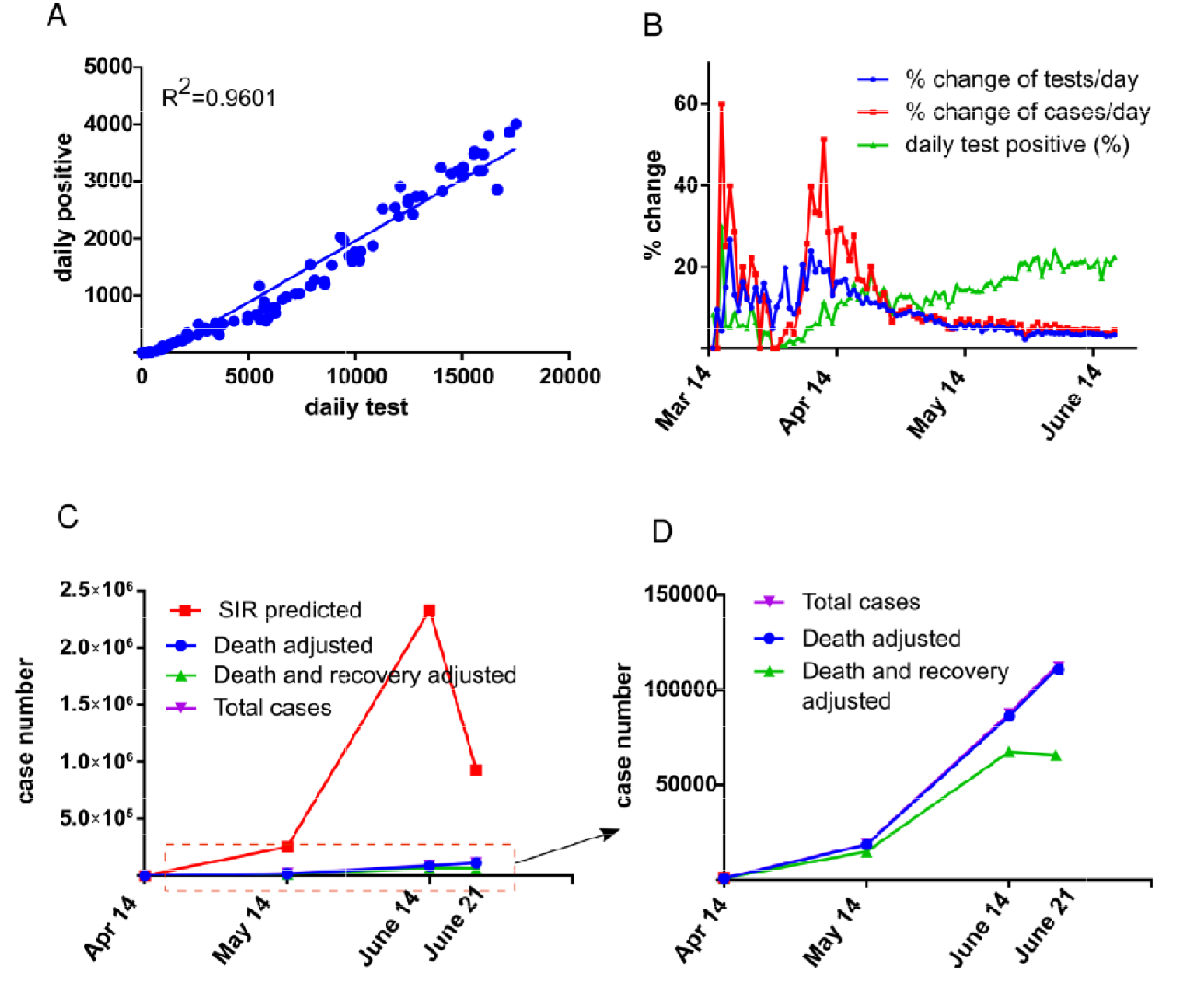
Real and SIR model data comparison. (A), daily test vs. daily positive case with linear regression fit (R^2^=0.9601); (B), daily percent change in test number and daily percent change in new cases are plotted with daily test positive (%) and (C), SIR predicted cases, real total cases, death adjusted cases and death with recovery adjusted cases were plotted in three monthly interval data point, and (D), part of Fig 2C (red dot rectangle) was further zoomed in to get better view. GraphPad Prism v. 6.0 was used to plot graphs and compiled in Inkscape v. 0.92.

We also selected three monthly time points started from March 14 to compare SIR predicted data with real cases, death adjusted cases and death with recovery adjusted cases (Figure 2C). As real data points are way below the predicted value we further zoomed the area to see trend of real data (Figure 2D). As SIR is close compartment model with death and recovery adjusted, we also subtracted death and recovery from total cases to get similar data for comparison. It showed that, from mid June death and recovery adjusted data is heading downward.

## 4 Discussion

### 4.1 Possible factors behind COVID-19 incidence pattern in Bangladesh

#### 4.1.1 Test number is a critical limiting factor

To understand the prevalence and distribution pattern of COVID-19, number of tests performed by a country plays critical role. Without enough testing true number of cases cannot be obtained. In addition, sufficient testing is required to properly declare COVID-19 related deaths and recovery. In our previous report (May 26, 2020), analyzing 91 countries’ number of tests performed per 100 K population data we found positive as well as negative correlation with positive cases and test positive (%) respectively with statistical significance [12]. Countries where limited numbers of tests were performed test positive (%) was comparatively higher. We reported that at that point global mean of test positive (%) was 9.94 ± 1.25 and countries with high number of tests (>500 tests per 100 K population) had test positive value well below 10% and countries with fewer tests (<500 tests per 100 K population) had average test positive rate of about 20% with extreme individual value of 85.51% [11]. As of 21 June, global mean of test positive (%) is 8.47±0.88. Countries with test numbers ranging from 500 to 5 K per 1 M population have average of 13.44 % test positives whereas countries with test number of above 50 K per 1 M population has test positive (%) value below 5. Bangladesh with 3,736 tests per 1 M population, monthly data showed that daily test positive (%) was increasing with declining daily percent increase in test numbers since beginning of May (Figure 2B). After reaching test positive rate around 20% the rate remained stable from early June onward. This indicated that initially the test positive (%) was getting higher due to very low number of tests performed. As most of the samples tested were either contact traced known of previous positive cases or people with very definite symptoms, test positive rate of 15-20% in Bangladesh was indicative of moderate to high infection in community. The pace of daily increase in testing capacity possibly could not follow the increasing infected patient numbers thus test positive (%) tend to rise. However, the stability of test positive (%) from beginning of June is indicative of saturated test positive numbers at current test capacity. However, the exact reason behind this could be multi dimensional.

To better understand the relationship we tabulated number of tests, test positive (%) and population density per km^2^ of some representative countries (Table 1). It is evident that with increasing test capacity, test positive rate decreased. One exception here is Sweden where official lock down or social distancing were not in place. It was also observed that although population density of Sweden is low, due to poor social distancing the virus could infect people very easily. Rather than direct effect of population density on test positive rate, it was more important at what extent direct person to person contact were restricted.

**Table 1:** Representative comparison of tests per 1M population, test positive (%) and population density per km^2^ of some countries along with Bangladesh

| Country | Tests/1M population | Test Positive (%) | Population Density/km<sup>2</sup> |
| --- | --- | --- | --- |
| Netherlands | 30,192 | 9.59 | 511 |
| Sweden | 38,193 | 14.53 | 25 |
| Italy | 82,436 | 4.78 | 205 |
| USA | 86,090 | 8.27 | 35 |
| Singapore | 98,508 | 7.31 | 7953 |
| Spain | 110,426 | 5.68 | 93 |
| UK | 116,241 | 3.86 | 274 |
| Russia | 116,481 | 3.44 | 8.82 |
| UAE | 307,270 | 1.48 | 135 |
| Bangladesh | 3,642 | 18.14 | 1239 |

[Data retrieved from Worldometers on June 22, 2020]

In the initial stage, Bangladesh Institute of Epidemiology, Disease Control and Research (IEDCR) alone collected and performed PCR test of suspected samples. Gradually test capacity was extended to public and private hospitals, universities, research organizations around the country which is at present more than 50 in number. Bangladesh is one of the low investing countries in health sector (health expenditure of 36.28 USD per capita) [13], thus the extension of test center and recruitment and training of expertise was relatively slow and still way below the need compared to large number of populations. Especially with the increasing rate of infection this can be vital limiting factor to know real case scenario in Bangladesh. These extended centers however are mainly in divisional headquarters or in large cities, where rural people staying away from the facility had difficulty to test as inter-city transport was closed. This has resulted some samples to become non usable and some false negative as samples might have been deteriorated due to longer transport and improper sample collection. We also identified that due to social non cooperation toward the COVID-19 positive or health care practitioners, many suspected individual were reluctant to give sample due to social fear. Although government hot line is open for all, many people had dissatisfactory experience calling for sample collection. Lack of management, long queue, waiting time in sample collection and test centers and delayed result also had negative impact, fear and rejection tendency among test seekers. These factors altogether had impact on uniform and broad data collection raising reliability issues on reported case numbers so far.

We further speculated possible incidence number as of 21 June if the test numbers in Bangladesh were close to high capacity testing countries (Table 2). To do this we calculated two possible scenarios where 50 K and 100 K tests per 1 M population were performed as of 21 June. If 5 to 10% test positives (%) were to be observed, approximately 0.4 to 0.8 million incidences could have been recorded. To achieve this Bangladesh had to perform around 13 times more tests compared to the tests done at present moment. This indicated that as Bangladesh are performing very low number of tests, it is hardly possible to achieve trend curve reaching peak and declining even though that amount of positives are present in community. It is also worth mentioning that limited test number can strongly influence the confirmation of death due to COVID-19 affecting true death count. It also can affect documentation of recovery numbers due to lack of subsequent testing of all confirmed positives.

**Table 2:** Tests per 1 M in Bangladesh as of 21 June and probable outcome if tests were performed at higher rate

| Test/1M | Number of tests | Positive case | Test positive (%) |
| --- | --- | --- | --- |
| 3736 | 6,15,164 | 1,12,306 | 18.26 |
| Probable tests/1M and approximate outcomes |  |  |  |
| 50,000 | 82,32,922 | 8,23,292 | if 10 |
|  |  | 4,11,646 | if 5 |
| 100,000 | 1,64,65,845 | 8,23,292 | if 5 |

#### 4.1.2 Population median age and obesity prevalence

In our previous report we also found that countries with high median age showed strong association with COVID-19 case fatality rate [12]. This was mainly due to prevalence of non communicable diseases as contributing factor for comorbidity and reduced immunity of elderly people. Bangladesh has median age of 27.90 years [14], with less elderly people. This could be one reason that young age group being dominant in the society are the most infected among all (55% belong to 21 to 40 years age group) [15]. Their inherent strong immunity might be a reason for silent infection and recovery with mild or even no symptoms at all. As of 31 May 2020 case fatality rate of Bangladesh is 1.38% which is below the global average of 4.26 ± 0.38 which could be due to less proportion of elderly in Bangladesh compared to the countries with high median age where case fatality climbed above 10%. Prevalence of obesity is linked to non communicable diseases which is low in Bangladeshi adult population (3.6%) [16]. Thus low obesity prevalence might have helped to minimize the co morbidity related complexity in Bangladeshi COVID-19 infected people.

#### 4.1.3 Environmental parameters

Scientists are having a wave of optimism that warmer weather might improve COVID-19 scenario [17]. Temperature, absolute humidity, relative humidity (RH), sunlight, ambient air flow, and altitudes were mostly studied parameters [18,19]. Respiratory viruses usually follow seasonal pattern, preferring either winter or summer, whereas some of them prefer to be year around virus [20]. Information regarding SARS-CoV-2 is still insufficient to label its seasonality; however, known other human corona viruses had shown clear preferences to winter. COVID-19 data so far showed temperate regions as the prevalence hot zone, but tropical areas are not completely out of the list. In one particular season incidence peak or hot spot may vary from virus to virus and usually avoid overlapping. Temperature and humidity determine the route of transmission and viral stability both in indoor and outdoor settings. Cold and dry weather dominate transmission via aerosol and small droplets whereas hot and humid weather facilitate transmission via fomites. Temperature above 30°C showed to block aerosol mediated Influenza virus transmission at variable RH, but contact mode transmission was still possible [21]. Surface stability and viability of SARS-CoV was shown to be lost at high temperature and high RH [22]. Bangladesh with monthly high temperature above 30°C and average temperature above 25°C from March to October may have reduced aerosol mediated viral transmission slowing community level infection rate.

UV index and air pollution may also play contributing role in viral transmission. Anthropogenic pollutants and microbes share common mechanism to confer immune deficiency [23]. These pollutants prepare the ground for COVID-19 like pandemic and further worsen the outbreak. Air particulate materials were found to be positively associated with COVID-19 incidence. Due to government imposed travel ban, reduced industrial effect air particulate emission was also low for the last two months which might have helped to reduce particulate material mediated virus transmission. Bangladesh has very high UV index during summer months with average of 10 or above with increased daylight hours [24]. UV index and daylight is related to ozone concentration which was found to reduce viability of viruses and COVID-19 transmission in a study conducted in Chinese cities [25]. In Bangladesh, ozone level in summer daylight may have helped to reduce viability of the viruses in environment [26]. Sunlight exposure which is also related to vitamin D production and immunity had helped to provide population level immunity to fight COVID-19 in Bangladesh [27]. Besides, with official leave, many people left capital city to stay with families in rural areas, where factors like low population density, air velocity, and sunlight exposure were helpful to restrict virus transmission.

#### 4.1.4 Heterologous immunity and food habit

Heterologous immunity, a form of cross reactivity is acquired from previously challenged unrelated microorganisms providing wider vaccine induced effectiveness and natural immunity against new infections [28,29]. Memory CD8+ T cells can help to detect newly infected viruses, but due to strain variation in RNA viruses less effective immunity is observed in human and in some cases show immune pathology[30]. Bangladesh is a densely populated country and people are exposed to different microorganisms during their lifetime, thus highly likely acquire heterologous immunity from natural infections. One study with cholera vaccination in Bangladesh showed natural immunity were long lasting compared to the oral vaccine [31]. In another study, Bangladeshi children were found to express more effector T cell activity compared to American children, supporting the ‘hygiene hypothesis’ [32]. According to this hypothesis early exposure to infectious agents may provide better immunity and in contrast lack of exposure may lead to allergies and autoimmune diseases. This also explains in part, reduced COVID-19 incidence in Bangladesh could be due to childhood acquired immunity against a variety of organisms. Co-infection of viruses is another aspect where one or more viruses may compete with another minimizing the virulence of the other [33]. Viral co-infection was reported in COVID-19 [34,35] and in Bangladesh as viral co-infection was observed in case of other viruses [36], co-infection of other respiratory viruses may have inhibitory effect on COVID-19 transmission and virulence. Food which boosts immune systems e.g. seasonal fruits, vegetables and spices could also play a positive role in slowing COVID-19 in Bangladesh.

## 5 Conclusion and outlook

Although many factors seemed to help Bangladesh to an extent, other factors like population density, urban population percentage and negligence toward health precautions still pose high risk of person to person disease transmission especially when offices and transport will be open. Climatic parameters may slow down COVID-19 transmission pattern and duration, however considering the size of pandemic its effect is modest [37]. Countries in both temperate and tropical regions must prepare for the possibility of severe outbreak; however, climatic variation will help to determine the local endemic cycle and seasonal peak. Step by step precautions are necessary to reduce sudden spike in infection number. Coming winter may face a second wave, so extended measures should be in place to contain the COVID-19 in Bangladesh. Public health policy and strategies need to be carefully adjusted considering these aspects to slow down the pandemic pace so that effective medical facilities can be provided to maximum possible people using available resources.

## Data Availability

Secondary data used and cited in this study are publicly available. Primary data will be available for researcher with proper citation.

